# Virtual Exam for Parkinson’s Disease Enables Frequent and Reliable Remote Measurements of Motor Function

**DOI:** 10.1101/2021.12.22.21268260

**Authors:** Maximilien Burq, Erin Rainaldi, King Chung Ho, Chen Chen, Bastiaan R. Bloem, Luc J.W. Evers, Rick C. Helmich, Lance Myers, William Marks, Ritu Kapur

**Author notes:** Correspondence to: Maximilien Burq, 269 E Grand Ave., South San Francisco, CA 94080, United States.

## Abstract

Sensor-based remote monitoring could help us better track Parkinson’s disease (PD) progression, and measure patients’ response to putative disease-modifying therapeutic interventions. To be useful, the remotely-collected measurements should be valid, reliable and sensitive to change, and people with PD must engage with the technology.

We developed a smartwatch-based active assessment that enables unsupervised measurement of motor signs of PD. 388 study participants with early-stage PD (Personalized Parkinson Project, 64% men, average age 63 years) wore a smartwatch for a median of 390 days, allowing for continuous passive monitoring. Participants performed unsupervised motor tasks both in the clinic (once) and remotely (twice weekly for one year). Dropout rate was 2% at the end of follow-up. Median wear-time was 21.1 hours/day, and 59% of per-protocol remote assessments were completed.

In-clinic performance of the virtual exam verified that most participants correctly followed watch-based instructions. Analytical validation was established for in-clinic measurements, which showed moderate-to-strong correlations with consensus MDS-UPDRS Part III ratings for rest tremor (ρ=0.70), bradykinesia (ρ=-0.62), and gait (ρ=-0.46). Test-retest reliability of remote measurements, aggregated monthly, was good-to-excellent (ICC: 0.75 - 0.96). Remote measurements were sensitive to the known effects of dopaminergic medication (on vs off Cohen’s d: 0.19 - 0.54). Of note, in-clinic assessments often did not reflect the patients’ typical status at home.

This demonstrates the feasibility of using smartwatch-based unsupervised active tests, and establishes the analytical validity of associated digital measurements. Weekly measurements can create a more complete picture of patient functioning by providing a real-life distribution of disease severity, as it fluctuates over time. Sensitivity to medication-induced change, together with the improvement in test-retest reliability from temporal aggregation implies that these methods could help reduce sample sizes needed to demonstrate a response to therapeutic intervention or disease progression.

## Introduction

Parkinson’s disease (PD) affects 6 million people world-wide as of 2018 - a number that is projected to grow to 12 million by 2040.^1^ Treatments are being developed to slow down or even halt the progression of PD.^2,3^ However, currently used endpoints (e.g. the MDS-UPDRS) exhibit high within-subject variability, and low test-retest reliability, which leads to inefficient clinical trials, and risks potentially missing relevant effects.^4^ Compounding this challenge, clinic-based physical exams provide only a snapshot of PD signs, and may not adequately reflect a patient’s functioning at home.^4,5^ Additionally, many people live far from major medical centers,^6^ so access to clinical trials of new therapeutics becomes restricted to a limited portion of the Parkinson population.^7,8^

These challenges have motivated the search for digital endpoints using wearable sensors, which allow for objective, frequent, and ecologically valid measurements of motor functioning in the patient’s home environment. Sensor-based remote monitoring could also help increase representation for groups whose data have historically not been included in clinical trials.^9,10^ Before such measurements can be used as endpoints in clinical trials to quantify disease progression, a careful evaluation of the clinical validity, reliability and sensitivity to change is required.^11,12^

A substantial volume of research has demonstrated the feasibility of using sensors placed on various parts of the body to quantify motor signs of PD.^13^ Results suggest that features extracted by digital signal processing can be correlated with clinical outcomes of interest, at least when tests are delivered in a controlled setting and assessments are supervised by a clinician.^14–22^ Active assessments measure patients’ maximum capacity, and can be complementary to passive monitoring, which measures the expression of signs in real life. Though some studies have probed the feasibility of using wearable sensors or smartphones for remote, self-guided active assessments, long-term engagement - which is critical to study disease progression - has been an important challenge.^23–25^ Studies focusing on passive monitoring of PD motor signs have generally not been able to capture a person’s intent to move, which is particularly relevant for signs of bradykinesia.^26^ Finally, test-retest reliability and sensitivity to clinically meaningful change have rarely been reported, and generally not on a large scale.

The smartwatch-based Parkinson’s Disease Virtual Motor Exam (PD-VME) can be deployed to remotely measure the severity of tremor, bradykinesia and gait impairment, via a self-guided active assessment.^27^ Here, we evaluate the feasibility of use and quality of data collected by the system, and report on the reliability, validity, and sensitivity to change of a set of digital measures derived from the PD-VME during a multi-year deployment in the Personalized Parkinson Project (PPP)^27^.

## Materials and methods

### Study design

Data were collected as part of the ongoing Personalized Parkinson Project (PPP), a prospective, longitudinal, single-center study (Clinical Trials NCT033648) of 520 people with early-stage Parkinson’s disease - diagnosed within the last 5 years.^27^ Study participants wear a smartwatch (Verily Study Watch) for up to 23 hours/day for the 3-year duration of the study, which passively collects raw sensor data from IMU, gyroscope, photoplethysmography and skin conductance sensors. All sensor data collection in this study used a wrist-worn wearable device, the 2nd-generation of the Verily Study Watch (hardware and firmware characteristics can be found in the supplementary materials).

Sensor data is collected during the yearly in-clinic MDS-UPDRS Part III motor exams. These are conducted both the on and the off state, after overnight withdrawal of dopaminergic medication (at least 12 hours after the last intake). Exams are video-recorded for quality controls and offline consensus scoring. Set 1 (N=198 participants) was selected for video-based consensus scoring by matching age, gender and MDS-UPDRS III score to be representative of the overall PPP study. Two assessors independently scored videos of the exams. When difficulties in rating MDS-UPDRS Part III tasks arose due to poor video quality, assessors provided scores only when confident in their assessment. MDS-UPDRS Part III consensus scores were computed as the median of the in-person rating and both video ratings.

Starting in May 2020, participants were offered the opportunity to enroll in a substudy, which asks them to perform an active assessment (Parkinson’s Disease Virtual Motor Exam, PD-VME) in the clinic and in remote, unsupervised settings. The PD-VME was deployed fully remotely, using digital instructions and an over-the-air firmware update to the watches of consented participants. 370 participants enrolled in the substudy (Set 2).

The smartwatch guides participants through the series of structured motor tasks comprising the PD-VME. It also allows patients on symptomatic medication to log the timing of their medication intake. The study design and patient-facing UI of the PD-VME are summarized in figure 1.

**Fig. 1.**
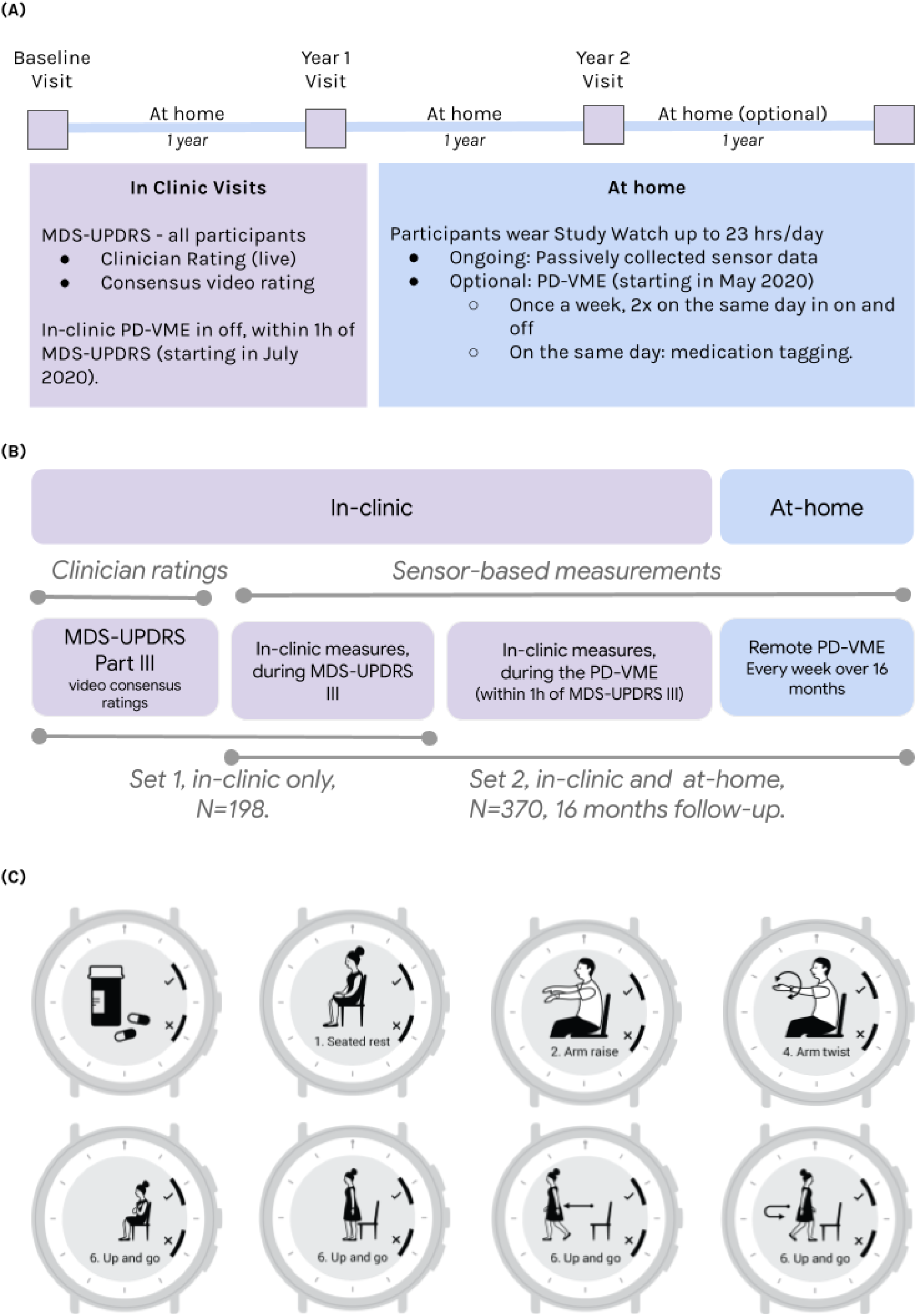
Study design and participant-facing interface: **(A)** Study design for the Personalized Parkinson’s Project. **(B)** Study schematic defining Sets 1 and 2 for the PD VME substudy. Set 1 consists of participants for whom consensus ratings of the MDS-UPDRS were available. Set 2 consists of participants who opted into the PD-VME substudy and performed at least one PD-VME. **(C)** User interface to prompt medication logging and PD-VME tasks. Seated rest, arm raise and arm twist (20s duration), up-and-go (60s duration).

Each week, participants were asked to perform the PD-VME twice on the same day, at two predefined times: first in the off state (selected as a time when they typically experienced their worst motor function), and then in the on state (at a time when they typically experienced good motor function later in the day). Participants not taking medication were instructed to complete the PD-VME twice, one hour apart. The helpdesk at the site (Radboudumc) monitored wear-time and PD-VME completion and reached out to participants if more than three consecutive weekly assessments were missed.

Starting in July 2020, participants enrolled in the PD-VME substudy were asked to perform the PD-VME during their in-clinic visit (in the same manner as they did remotely), while the assessor observed its execution without providing feedback or any additional instructions. The in-clinic PD-VME is performed within 1 hour after completion of the MDS-UPDRS part III off state exam, and before dopaminergic medication intake.

### Ethics

The study is being conducted in conformance with the Good Clinical Practice ICH E6 guideline and in compliance with the Ethical Principles for Medical Research Involving Human Subjects, as defined in the Declaration of Helsinki (version amended in October 2013), the Dutch Personal Data Protection Act, and the European General Data Protection Regulation. The Commissie Mensgebonden Onderzoek Region Arnhem-Nijmegen approved the study protocol and communication materials.^28^ Signed informed consent is obtained before engaging a participant in any study procedure, including use of the PD-VME.

### Design of the Parkinson’s disease virtual motor exam

The PD-VME system, including participant-facing training materials, user interface, task choice and digital measures, was developed using a patient-centric approach.^28^ The PD-VME comprises eight tasks designed to assess various domains of motor signs: rest and postural tremor, upper extremity bradykinesia through finger tapping, pronation-supination and repeated hand opening and closing, lower-extremity bradykinesia through foot stomping, gait and postural sway. Participant-facing instructions for each task are in Supplementary Table 1. Figure 1.C shows the PD-VME user interface for the four tasks analyzed in this paper. Selection of targeted signs was informed by research on meaningful aspects of health in PD: tremor, bradykinesia and gait were identified as three of the top four symptoms that people with PD most want to improve.^29^ A patient panel of PPP participants was involved throughout the design process to assess and improve the usability of the system.

**Table 1.** Participant demographics and characteristics at their first clinic visit.

|  |  | Set 1 (consensus)<sup>a</sup> | Set 2 (PD-VME)<sup>b</sup> |
| --- | --- | --- | --- |
| **Demographics** |  |  |  |
| N<sup>c</sup> |  | 198 | 370 |
| Age (years) | Mean (SD) | 62.9 (8.9) | 62.3 (8.8) |
| Gender | Men, n (%) | 128 (64.6) | 231 (62.4) |
| Side on which the watch was worn | Left, n (%) | 182 (92.0) | 342 (92.4) |
| Side on which the watch was worn | Most affected, n (%) | 117 (59.1) | 196 (53.0) |
| Disease at onset |  |  |  |
| Time since diagnosis (years) | Mean (SD) | 3.3 (1.4) | 2.9 (1.4) |
| **Disease severity** |  |  |  |
| MDS-UPDRS Part I | Mean (SD) | 15.7 (8.3) | 15.1 (7.7) |
| MDS-UPDRS Part II | Mean (SD) | 9.6 (6.4) | 8.4 (5.7) |
| MDS-UPDRS Part III (Off state) | Mean (SD) | 37.6 (15.3) | 33.8 (13.0) |
| Hoehn and Yahr Scale | Mean (SD) | 2.1 (0.6) | 2.0 (0.5) |
| **Medications** |  |  |  |
| Participants on any symptomatic medication | n (%) | 185 (93%) | 297 (80%) |
| Participants on fast-acting medication<sup>d</sup> | n, (%) | 117 (59%) | 202 (54%) |

<sup>a</sup>Set 1 (consensus) includes all participants who had at least two independent video ratings and one in-clinic rating. <sup>b</sup>Set 2 (PD-VME) includes all participants who had at least one PD-VME exam. <sup>c</sup>180 participants were included in both sets, for a total of 388 unique participants. <sup>d</sup>excludes dopamine agonists.

During execution of PD-VME tasks, tri-axial accelerometer and gyroscope data was collected at a sample rate of 200 Hz. For each task, an initial list of concepts of interest were identified (e.g. tremor severity, quality of gait). For each concept, digital signal processing was implemented to convert the raw sensor data into 11 exploratory outcome measures (e.g. tremor acceleration, arm-swing magnitude)

### Evaluation of digital measures from the PD-VME

Participant engagement with the PD-VME, measured as the fraction of participants who performed at least one complete exam in a given week, was evaluated over the course of 70 weeks. The ability of the participants to perform the PD-VME correctly without having received in-person instructions was assessed using the assessor observations from the in-clinic PD-VME.

The analytical validity, reliability, and sensitivity to change of digital measurements from the PD-VME was evaluated. First, the analytical validity of measures, collected during the in-clinic MDS-UPDRS, was assessed using the Spearman correlation coefficient of the measure against the consensus of three independent MDS-UPDRS Part III clinical ratings. Second, the test-retest reliability in the home setting was evaluated by computing the intra-class correlation between monthly means across subsequent months for months with no missing PD-VME. Finally, the sensitivity to change was assessed by testing the ability of the remote measurements to distinguish between the off and the on states for the subset of patients in Set 2 who are on dopaminergic medication. An unsupervised PD-VME exam is determined to be in the off state if it occurred at the pre-scheduled off time *and* at least 6 hours after a medication tag. Similarly, an exam is determined to be in the on state if it occurred at the pre-scheduled on time *and* between 0.5 and 4 hours after a medication tag. Two measures were used to assess the magnitude of change: mean difference (and associated 95% confidence interval) and Cohen’s d. Participants taking dopamine agonists were not included in the on-off comparison because of their prolonged effect.

For each task, one outcome measure is shown in the results, selected on the basis of its high performance across all three aspects (analytical validity, test-retest reliability and sensitivity to change) for inclusion in the results. The results of the remaining 8 measures are presented in the supplementary materials.

### Comparison of the in-clinic and remotely collected PD-VME measurements

To characterize the extent to which measures obtained from clinic-based physical exams (off) reflected patients’ signs in the remote setting (off), we compared the distributions of participants’ in-clinic and remote PD-VME outcomes (completed within 90 days of the clinic visit). A subset of N=194 participants from Set 2 who performed the PD-VME in-clinic were included in this analysis.

Figures and statistical analyses were generated using the Python programming language, using the SciPy^30^, Matplotlib^31^ and seaborn^32^ libraries. In all numerical results that follow, point estimates are followed by 95% confidence intervals in square brackets. Confidence intervals were calculated using the bootstrap method with 1000 resampling iterations.

## Results

Demographic and clinical characteristics of the study population are presented in Table 1, for participants in Set 1 and Set 2.

### Engagement

Median smartwatch wear time across all PPP participants (N=520)^33,27^ was 22.1 hours/day, with a median follow-up period of 390 days. Variations in follow-up duration are due largely to the N=126 who have not completed the study at the time of publication, and loss-to-follow-up is only 2%. Participants in Set 2 completed 22,668 PD-VMEs, corresponding to 59% of per-protocol test sessions during the 70-week follow-up period (Supplementary figure 1). 80% of participants had at least 1 PD-VME in the first week, and 40% had completed one PD-VME in week 52.

### Useability

Participants’ ability to perform the PD-VME was assessed during the in-clinic visit. Participants were able to complete the tasks in the exam (100% for tremor and upper-extremity bradykinesia and 98.5% for gait). Major protocol deviations were recorded as follows: 8.2% of participants did not place their hands on their lap during rest tremor tasks, 3.1% of participants performed the arm-twist using both arms in 3.1% and 6.8% of participants either walked with their arms crossed across their chest or sat down repeatedly during the gait task. Detailed results are summarized in supplementary table 2.

### Rest tremor

Among three measurements that were considered for measuring tremor severity, lateral tremor acceleration measurement is presented here because it showed the strongest correlation to in-clinic MDS-UPDRS ratings, and the strongest ability to separate on from off state measurements. Results for additional measures are included in Supplementary Table 2.

The Spearman rank correlation between the median lateral acceleration during the rest tremor task and expert consensus rating of MDS-UPDRS task 3.17 was 0.70 [0.61, 0.78], N=138 (Fig. 2 panel A). 56 participants did not have sufficient video quality to ensure high confidence consensus ratings Wrist acceleration signals intuitively map to the clinical observations during the MDS-UPDRS (figure 2.B).Next, the sensitivity to on-off changes of the rest-tremor acceleration measurement was assessed (Fig. 2.C). A small effect (Cohen’s d of 0.2) was observed comparing the on and off state. The mean difference in the measure was 0.10 [0.06, 0.14].

**Fig. 2.**
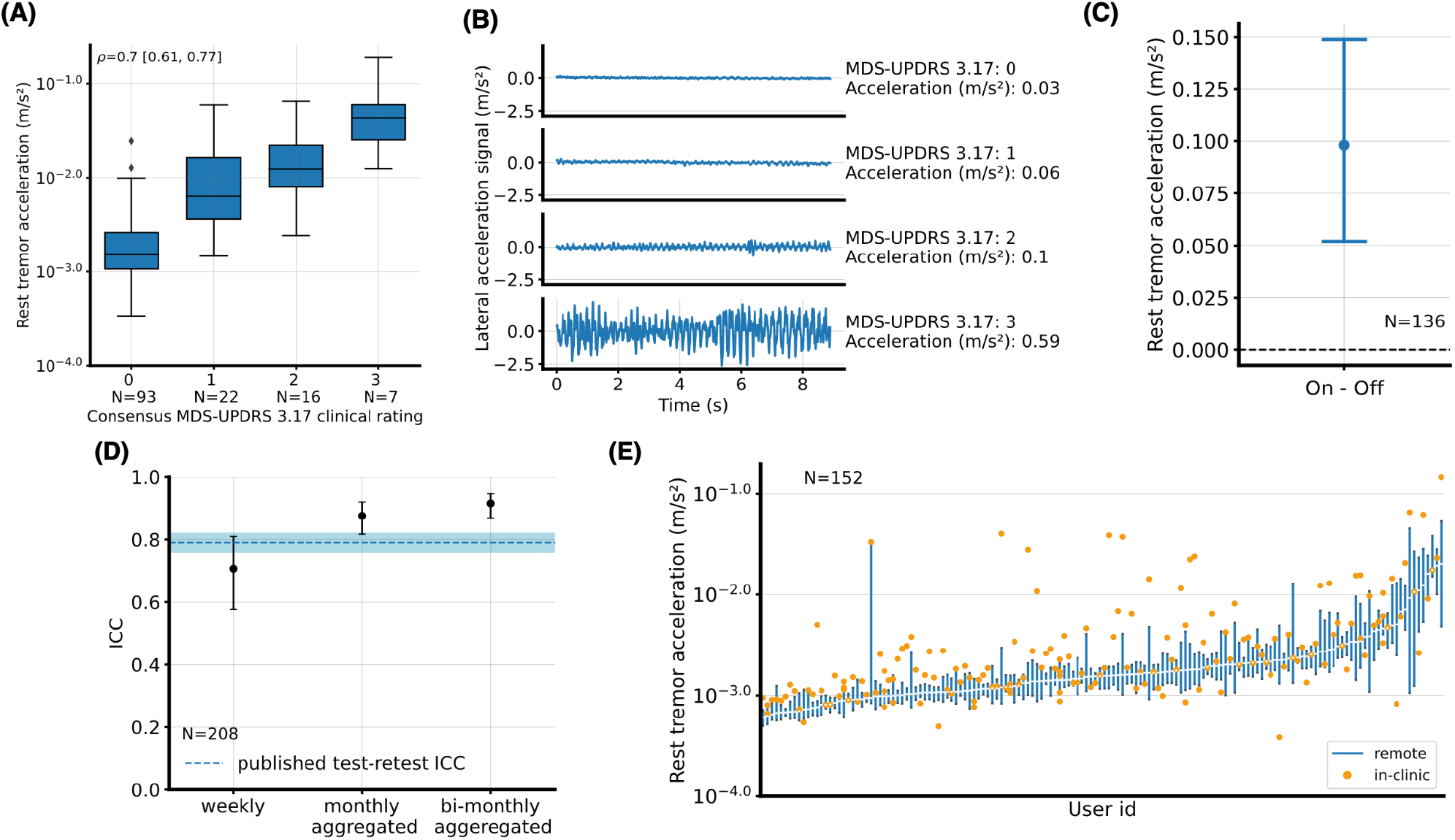
Rest tremor acceleration. **(A)** Lateral tremor acceleration (log scale), measured during the in-clinic examination, by rest tremor (MDS-UPDRS 3.17) consensus score. **(B)** Illustrative examples of raw lateral acceleration signals for each score on the MDS-UPDRS 3.17. Measurement values, as computed by the PD-VME, are also indicated. **(C)** Difference between the remote measurements in on and off states, aggregated over PD-VMEs obtained during the first two months from each participant. Mean and 95% confidence intervals across participants are represented. **(D)** Intra-class correlation (ICC) between at-home measurements, for various durations of aggregation. Whiskers represent 95% confidence intervals. The dotted blue line represents the published test-retest ICC of 0.79 for the whole rest tremor UPDRS Part III subcomponent (all four extremities + lip & jaw).^34^ **(E)** Distribution of PD-VME tremor measurements (off state) obtained during the in-clinic PD-VME (orange dot, representing a single measurement) and remote PD-VMEs (blue bar, representing the 25th to 75th percentile of PD-VMEs within 90 days of the in-clinic PD-VME), sorted on the remote PD-VMEs.

Test-retest reliability is reported in figure 2.D, with intra-class correlation (ICC) of 0.71 [0.58-0.81] week-on-week (N=208), and ICC of 0.90 [0.84-0.94] m.s^-2^ for monthly averaged measures (N=139).

Finally, the distribution of remote measurements compared to the sensor measurement during the in-clinic VME is shown in figure 2.E. The in-clinic PD-VME measure was between the 25th and the 75th percentiles of the remote PD-VME measures for 41% of the participants.

### Upper-extremity bradykinesia

Among the four measurements that were considered for measuring upper-extremity bradykinesia severity, no single measure showed both strong correlation to in-clinic MDS-UPDRS ratings, and a strong ability to separate on from off state measurements. Therefore, results are included below for both the arm-twist amplitude, and the arm-twist rate.

The highest correlation with expert consensus rating of MDS-UPDRS task 3.6 was observed for the arm twist amplitude measure, with ρ = -0.62 [-0.73, -0.50], N=159 (Fig. 3.A). However, the effect of medication state (Cohen’s d of -0.07) was very small (Fig. 3.C).^35^ The mean on-off difference in the measure was -0.9 [0.0, -1.6] degrees. Test-retest ICC (figure 3.D) was 0.71 [0.59-0.80] week-on-week (N=208) and 0.89 [0.84-0.94] for monthly-averaged measures (N=136). The in-clinic PD-VME measure was between the 25th and the 75th percentiles of the remote PD-VME measures for 45% of the participants.

**Fig. 3.**
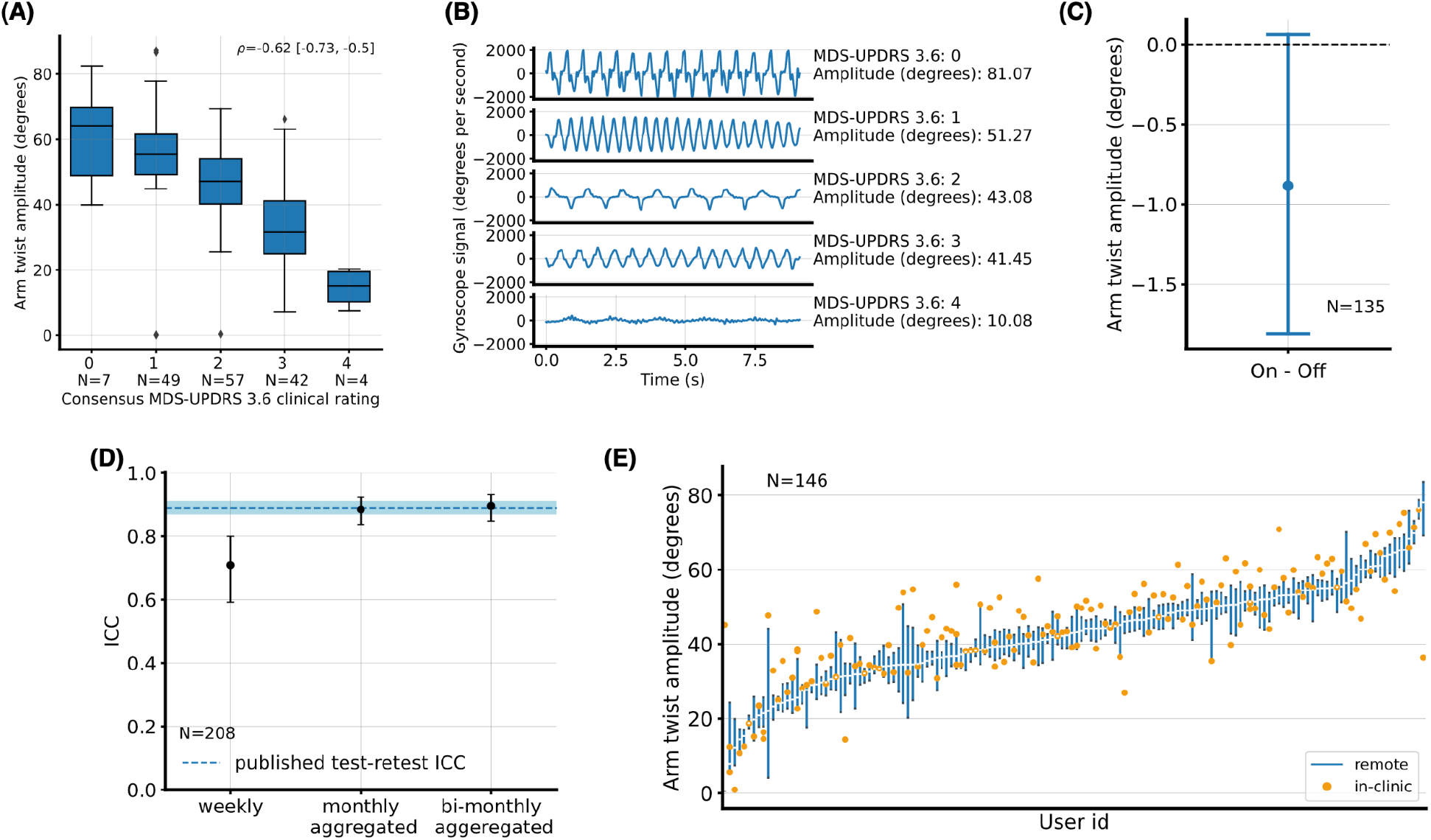
Upper extremity bradykinesia. **(A)** Arm twist amplitude measured during the in-clinic examination, by pronation-supination (MDS-UPDRS 3.6) consensus scores. **(B)** Illustrative examples of raw gyroscope signals, along the x axis, for each score on the MDS-UPDRS 3.6. Measurement values, as computed by the PD-VME, are also indicated. **(C)** Difference between the remote measurements in on and off states, aggregated over PD-VMEs obtained during the first two months from each participant. Mean and 95% confidence intervals across participants are represented. **(D)** Intra-class correlation (ICC) between at-home measurements, for various durations of aggregation. Whiskers represent 95% confidence intervals. The dotted blue line represents the published test-retest ICC of 0.89 for the whole bradykinesia subcomponent of the UPDRS Part III.^31^ **(E)** Distribution of PD-VME arm-twist measurements (off state) obtained during the in-clinic PD-VME (orange dot, representing a single measurement) and remote PD-VMEs (blue bar, representing the 25th to 75th percentile of PD-VMEs within 90 days of the in-clinic PD-VME), sorted on the remote PD-VMEs.

The assessors observed during the in-clinic PD-VME exam that some patients mainly focussed on the speed of the arm-twist movement rather than the amplitude. Therefore, sensor-based measures of the rate of arm-twist and the combination of rate and amplitude were investigated as well. Correlations to the consensus MDS-UPDRS ratings of ρ = 0.06 [-0.25, +0.13] for arm-twist rate, and ρ = -0.42 [-0.55, -0.28] for the product of rate and amplitude were observed. Both metrics showed significant change in on and off: Cohen’s d of -0.22 and mean change of -0.16 [-0.13, -0.20] s^-1^ for arm-twist rate, and Cohen’s d of -0.26 and mean change of -8 [-6, -10] degrees/s for the combination. The full results are included in Supplementary Table 4.

### Arm swing during gait

Among the three measurements that were considered for measuring gait impairment, arm swing acceleration was selected. While it was not the best outcome measure across any of the criteria, it showed solid performance across all of them. Results for the measures that were not selected are included in Supplementary Table 5.

The Spearman rank correlation between the arm swing acceleration during the gait task and expert consensus rating of MDS-UPDRS task 3.10 was ρ = -0.46 [-0.57, -0.31], N=164 (Fig. 4.A). A small effect (Cohen’s d of 0.44) was observed comparing the on and off state. The mean difference in the measure was -0.8 [-1.2, -0.5] m.s^-2^. Test-retest ICC (figure 4.D) was 0.43 [0.30-0.56] week-on-week (N=210), and 0.75 [0.66-0.84] for monthly-averaged measures (N=139). The in-clinic PD-VME measure was between the 25th and the 75th percentiles of the remote PD-VME measures for 39% of the participants.

**Fig. 4.**
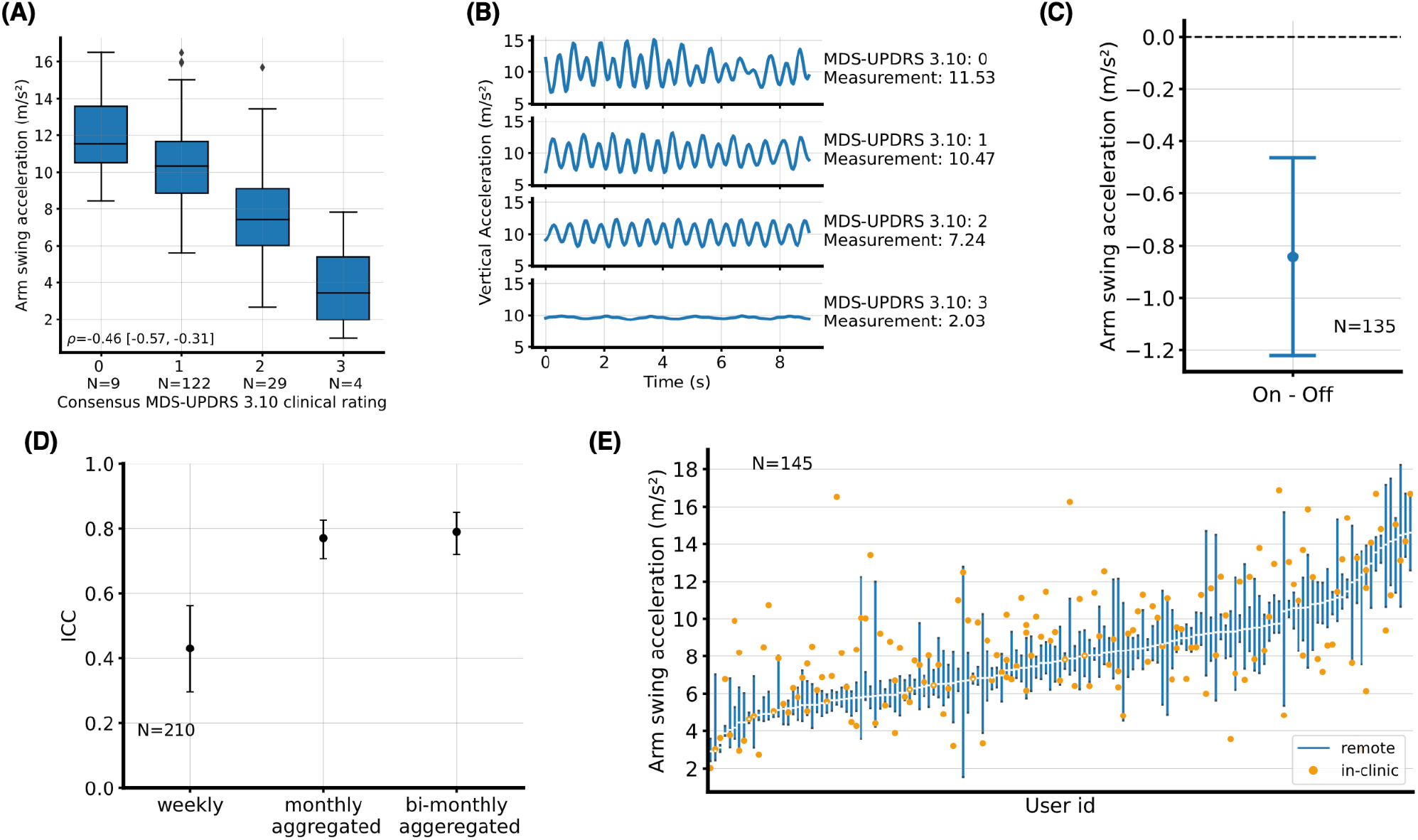
Gait. **(A)** Arm swing acceleration measured during the in-clinic examination, separated by gait (MDS-UPDRS 3.10) consensus scores. **(B)** Illustrative examples of raw accelerometer signals, for each score on the MDS-UPDRS 3.10. Measurement values, as computed by the PD-VME, are also indicated. **(C)** Difference between the remote measurements in on and off states, aggregated over PD-VMEs obtained during the first two months from each participant. Mean and 95% confidence intervals across participants are represented. **(D)** Intra-class correlation (ICC) between at-home measurements, for various durations of aggregation. Whiskers represent 95% confidence intervals.^34^ **(E)** Distribution of PD-VME gait measurements (off state) obtained during the in-clinic PD-VME (orange dot, representing a single measurement) and remote PD-VMEs (blue bar, representing the 25th to 75th percentile of PD-VMEs within 90 days of the in-clinic PD-VME), sorted on the at-home PD-VMEs.

## Discussion

The data from this study suggest that people with PD engage with and are able to use the PD-VME, and that the quality of data collected is high enough to enable evaluation of the analytical validity, reliability, and sensitivity to change of digital measures built from this system.

A digital solution is only useful if people with PD engage with it regularly. We observed robust levels of engagement, both in terms of overall wear time (>21 hours/day) and engagement with the active assessment, which was 59% over one year when assayed on a weekly basis. This is at the high end of reported values,^23,24^ and this suggests that combining active assessments with passive monitoring on smartwatch form-factors have the potential to yield substantial quantities of high quality data. For studies assessing longitudinal progression, our observations suggest that even higher engagement could be obtained by requiring a set of weekly unsupervised tests for a limited duration at baseline and again at the end of the follow-up period.

We showed a moderate-to-strong correlation between in-clinic MDS-UPDRS Part III measurements and consensus clinical ratings for rest tremor, bradykinesia, and arm swing during gait, which provided analytical validation of the individual measurements. These results are on par with similar published analyses of wrist-worn sensors ^36–39^ and demonstrate the ability of the PD-VME to provide metrics that map to the observations of an expert clinician. While the moderate-to-strong correlations with MDS-UPDRS scores establish that the measurements are working as intended, engineering for perfect correlation simply recreates an imperfect scoring system, and washes out the potential for increased sensitivity of sensor-based measurements. One key reason for making a shift towards digital assessments is that clinical scores remain subjective in nature, and use a low resolution, ordinal scoring system. The criteria for transitioning between different scores leave much room for subjective interpretation, and cause considerable variability between and within raters in daily practice.^4^

This is exemplified by the results shown for the upper-extremity bradykinesia measure, in which we find that the measure most correlated with in-clinic MDS-UPDRS ratings - amplitude of arm-twist - is not the one that is most sensitive to change from dopaminergic medication. It is possible that while the experts are instructed to evaluate “speed, amplitude, hesitations, halts and decrementing amplitude”,^40^ they may focus mostly on amplitude. Similarly, we observe a gradient of tremor measurements, both in-clinic and remotely, even within the group of participants who are rated as a 0 on the MDS-UPDRS 3.15 or 3.17. This suggests that some amount of tremor could be present, both in the clinic and at-home, even before it becomes apparent to the human eye. Indeed, it is generally a well-accepted phenomenon that tremors are easier felt or even heard (using a stethoscope) than observed by an examiner. This reinforces the need for objective sensor-based measures, and the need to evaluate these measures based on their ability to detect clinically meaningful changes rather than simply reproducing subjective clinical exams.

In people with PD, dopaminergic medication can considerably improve severity of motor signs over short time frames. This “on-off” difference is well-accepted as a clinically meaningful change, and when coupled with wearable sensors and patient-reported tagging of daily medication regimen, creates multiple “natural experiments” in the course of patients’ daily lives. These allow us to test the clinical validity ^41,42^ of the PD-VME measures as pharmacodynamic/response biomarkers for people with PD in the remote setting. Indeed, we demonstrate that digital measures for tremor, upper-extremity bradykinesia and gait are able to detect significant change in patients’ motor signs before and after medication intake.

For clinical trials aiming to show disease modification, measurements that provide reliable estimates of a subject’s disease state can increase statistical power, and reduce the required sample size or trial duration. However, measuring long-term progression using infrequent measurements is difficult, because motor and non-motor signs of PD can markedly fluctuate from moment to moment,^37,43^ depending on factors such as the timing of medication intake or the presence of external stressors. The increased test-retest reliability of the monthly aggregated measures from this study suggest that collecting outcome measures remotely and at an increased frequency increases their reliability, and have the potential to measure progression of the average motor sign severity.

This work is not without its limitations. The smartwatch was worn unilaterally, though PD typically exhibits asymmetrical symptom severity. Asymmetry may have particularly affected our assessments of gait which may exhibit different characteristics when observed from the most- or least-affected upper-extremity. Further analysis is needed to better understand the impact of device placement on measurement validity and reliability. Also, PD is multifaceted in nature, and signs manifest along multiple motor and non-motor domains.^44^ While we present data on multiple important motor domains, additional research is needed to use the rich data collected in this study to expand this to additional motor and non-motor aspects (e.g. by using the PPG and EDA data collected). Finally, future work replicating the results presented here in a different study population is needed. In addition to enabling exploratory analyses around non-motor symptoms, the 3-year follow-up of PPP will enable future work looking at the PD-VME’s sensitivity to long-term disease progression. The smart-watch form-factor could enable future analysis combining the PD-VME measurements together with measures obtained from passive monitoring. For signs of bradykinesia in particular, active tasks allow us to capture the intent to move, which may be difficult to capture from passively collected data.

In conclusion, these data suggest that patients engage robustly with the PD-VME, and are able to complete remote active assessments of motor function to yield data of a sufficient quality to generate digital measurements of motor signs, test their analytical validity, and assess their sensitivity to change in medication status. The system allows for an increased frequency of data collection, enabling monthly aggregation of measurements, leading to increased test-retest reliability. In turn, high reliability suggests that these measures have potential as digital biomarkers of progression. Further research is needed to more firmly establish the ability of these and other measures to serve as progression biomarkers.

## Data Availability

The datasets generated during this study will be made available to qualified researchers worldwide, insofar as such requests are for analysis for which the participants in the PPP have provided informed consent. Requests for access to data are governed by a Research and Data Sharing Review Committee (RDSRC), whose role is to protect subjects' privacy by limiting the availability of the study data and controlling access to sources of information that might potentially be used to identify the individual subjects associated with the biospecimen analyses. The RDSRC will assess the relevance and scientific quality of research proposals for which study data or material is requested.

## Abbreviations

ICC: Intra-class correlation
MDS-UPDRS: Movement Disorder Society Unified Parkinson’s disease Rating Scale
PD-VME: Parkinson’s Disease Virtual Motor Exam
PPP: Personalized Parkinson’s Project

## Author contributions

Conceptualization: MB, RK, WJM, BRB, ER, LM

Methodology: MB, RK, ER, BRB

Software: MB, KCH, ER

Validation: MB, KCH, CC, BRB

Formal Analysis: MB, KCH, ER, BRB

Investigation:, BRB, RK

Resources: BRB, RK

Data Curation: MB, ER, KCH, BRB

Writing – original draft: MB, ER, KCH, CC, RK

Writing – review & editing: BRB, WJM, LE, RH, LM

Visualization: KCH, MB, ER, RK

Funding acquisition: RK, WJM, BRB

Supervision: RK, LM, BRB

Project Administration: BRB, RK

## Acknowledgements

We are grateful to the Personalized Parkinson’s Project participants who shared their time interacting with the PD-VME, provided invaluable feedback on the design and user experience, and shared their experience with PD. For their contributions to the study design, study operations and data collection, we would like to recognize Cindy Yee, Anisha Choudhury, Kinjal Satasiya, Nicole Greco, Tessa van de Zande, Myrthe Burgler, Marjan Meinders, Sabine van Zundert, Debbie de Graaf, and Ginny Snellen. For reviewing and providing feedback on the manuscript, we would like to acknowledge: Nate Kowahl, Sooyoon Shin, Li-Fang Cheng, Niranjan Sridhar, Zachary Owen and Sara Popham.

## Data Availability

The datasets generated during this study will be made available to qualified researchers worldwide, insofar as such requests are for analysis for which the participants in the PPP have provided informed consent. Requests for access to data are governed by a Research and Data Sharing Review Committee (RDSRC), whose role is to protect subjects’ privacy by limiting the availability of the study data and controlling access to sources of information that might potentially be used to identify the individual subjects associated with the biospecimen analyses. The RDSRC will assess the relevance and scientific quality of research proposals for which study data or material is requested.^27^

## Code Availability

The computer code used in this study will be open-sourced and available for academic research purposes, alongside the study data, for qualified researchers worldwide.

