## Supplementary materials for "Virtual Exam for Parkinson’s Disease Enables Frequent and Reliable Remote Measurements of Motor Function"

#### Informed consent

The study is being conducted in conformance with the Good Clinical Practice ICH E6 guideline and in compliance with the Ethical Principles for Medical Research Involving Human Subjects, as defined in the Declaration of Helsinki (version amended in October 2013), the Dutch Personal Data Protection Act, and the European General Data Protection Regulation. The Commissie Mensgebonden Onderzoek Region Arnhem-Nijmegen approved the study protocol and communication materials.[28] Signed informed consent is obtained before engaging a participant in any study procedure, including use of the PD-VME.

#### Description of the hardware platform

All sensor data collection in this study used a wrist-worn wearable device, the 2nd-generation of the Verily Study Watch. It features an inertial measurement unit (IMU) (3-axis accelerometer with range +/- 16 G; and 3-axis gyroscope with range +/- 2000 DPS), photoplethysmography, electrocardiogram (ECG), and electrodermal activity sensors, and several environmental sensors. In this study, the IMU is sampled at 100 Hz, except during a MDS-UPDRS or PD-VME exam, during which it is sampled at 200 Hz. The device was worn on the participant's preferred side (see Table 1).

#### Participant-facing instructions for the PD-VME.

| Task | Instructions to the participant |
|---|---|
| Finger tapping on watch | Place the thumb of your other hand on the watch frame and then allow your fingers to gently grasp your Study Watch arm in a natural position. From here, extend your index finger so that it is hovering above the |

| | |
|---|---|
| | watch face, ready to tap.<br><br>Use your index finger to tap on the target as quickly and forcefully as possible until you feel a vibration and receive notification from the Study Watch when the task is complete in 20 seconds. |
| Rest tremor | Remain seated comfortably in a chair with your hands resting on your thighs, making sure to keep your palms up.<br><br>Close your eyes, and count back from 100 aloud until you feel a vibration and receive notification from the Study Watch when the task is complete in 20 seconds. |
| Postural tremor | Remain seated comfortably in a chair and hold both of your arms out in front of you, palms facing down.<br><br>Close your eyes, and count back from 100 aloud until you feel a vibration and receive notification from the Study Watch when the task is complete in 20 seconds. |
| Hand Open and Close | On the side where you’re wearing the Study Watch, hold your arm out in front of you with your hand up, palm facing away from you.<br><br>Open and close your hand as completely and quickly as possible. Repeat this movement until you feel a vibration and receive notification from the Study Watch when the task is complete in 20 seconds. |
| Arm Twist (Pronation/Supination) | Remain seated comfortably in a chair. On the side where you’re wearing the Study Watch, hold your arm out straight.<br><br>Rotate your hand, alternating between palm to the ceiling and palm facing the floor. Repeat this movement as completely and quickly as possible until you feel a vibration and receive notification from the Study Watch when the task is complete in 20 seconds. |
| Foot Stomp | Use the hand wearing the Study Watch to grasp your knee on the same side of your body.<br><br>Lift the leg you are grasping while continuing to hold onto your knee, and then stomp your heel on that side on the ground. Repeat this movement as completely and quickly as possible until you feel a vibration and receive notification from the Study Watch when the task is complete in 20 seconds. |
| Up-and-go | Sit in a chair and cross your arms in front of your chest, with your Study Watch arm on top of your other arm.<br><br>Stand up, pause briefly, then place your arms at your sides. Walk in a straight line at a natural, comfortable pace in your longest room, turning |

| | |
|---|---|
| | around and walking back and forth as needed. Repeat this movement until you feel a vibration and receive notification from the Study Watch when the task is complete in 60 seconds. |
| Stand Still (Postural Stability) | Stand upright. Cross your arms across your chest, with your Study Watch arm on top of your other arm.<br><br>Stand still while looking straight ahead until you feel a vibration and receive notification from the Study Watch when the task is complete in 60 seconds. |

**Supplementary table 1.** Participant-facing instructions for the VME.

### Engagement

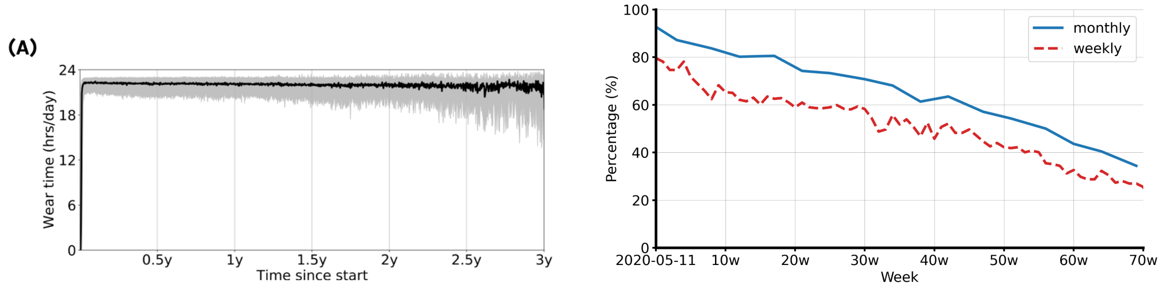


**Supplementary figure 2, Engagement: (A)** Study Watch median daily wear time (hours) by day of study across the whole PPP cohort. The grey shaded area represents the 25th and 75th percentiles of wear time for a given day. The median wear time is 22.1 hours per day. The number of participants used to compute the median wear time decreases with the increase of "time since start", because a proportion of participants have been enrolled <3 years ago and are still actively contributing data. **(B)** Percentage of participants in set 2 with at least one PD-VME per week.

### Observations of in-clinic performance of the PD-VME

**Supplementary table 2: Deviations in the execution of the in-clinic PD-VME, observed by the assessors (percentage followed by number of cases in brackets). For each participant (n=292), only the first in-clinic PD-VME after receiving remote instructions is included. a: missing because the participant did not perform this task during the visit (e.g. due to severe gait impairments in the off state).**

| PD-VME task | Any deviation observed | Observed major deviations | Observed minor deviations |
|---|---|---|---|
| Resting while sitting (rest tremor) | 46.6% (136)<br>Missing: 0% (0) | Hands not on lap: 27.4% (37) | - Participant does not count or does not count out loud<br>- Eyes not closed<br>- Self-rated tremor on the wrong side<br>- Hand palms facing down |
| Lifting arms (postural tremor) | 18.2% (53)<br>Missing: 0% (0) | Moved hands up and down: 1.4% (4) | - Participant does not count or does not count out loud<br>- Eyes not closed<br>- Self-rated tremor on the wrong side |
| Standing up | 17.8% (52)<br>Missing: 1.0% (3)[a] | Used hands when standing up: 3.4% (10) | - Difficulties rating both walking and standing at the same time<br>- Arms not crossed |
| Walking | 7.5% (22)<br>Missing: 1.4% (4)[a] | Crossed arms during walking: 3.4% (10)<br>Interrupted by sitting down: 3.4% (10) | - Difficulties rating both walking and standing at the same time. |
| Pronation supination | 5.1% (15)<br>Missing: 0% (0)[a] | Performed using both arms: 3.1% (9) | - Focus on either speed or amplitude (rather than both) |

Focus on either speed or amplitude (rather than both) among all 292 participants: 9.6% (28)

### Supplementary results: Tremor

Three methods for tremor severity algorithms were considered, based on published literature and authors' previous experience with similar wrist-worn devices: lateral tremor acceleration, total tremor acceleration, lateral tremor amplitude. Lateral tremor acceleration is obtained by computing the median absolute tremor acceleration along the lateral axis of the accelerometer signal, after filtering out the gravitational component. This is the axis parallel to 3 and 9 o'clock on the watch face. Total tremor acceleration is obtained similarly, but on the norm of the acceleration vector rather than a single axis.

The final algorithm was selected based on correlations with the in-clinic MDS-UPDRS rest tremor (3.17) single-rater score, for a subset of patients from Set 2 (some of which also belonged to Set 1). For patients who also belonged in Set 1, only clinic visits where consensus scores were not collected were used for algorithm selection.

| method | Spearman R vs clinical ratings | Test-retest ICC (month-on-month) | on_off separation (Cohen's D) |
|---|---|---|---|
| Lateral tremor acceleration (log) | 0.7 - [0.61; 0.78] | 0.96 - [0.93;0.98] | 0.20 |
| Lateral tremor amplitude (log) | 0.64 - [0.53; 0.72] | 0.94 - [0.91;0.96] | 0.14 |
| Total tremor acceleration (log) | 0.71 - [0.62; 0.78] | 0.95 - [0.92;0.97] | 0.16 |

**Supplementary Table 2.** Rest tremor methods.

| method | Spearman R vs clinical ratings | Test-retest ICC (month-on-month) | on_off separation (Cohen's D) |
|---|---|---|---|
| Lateral tremor acceleration (log) | 0.59 - [0.47; 0.69] | 0.94 - [0.9;0.97] | 0.19 |
| Lateral tremor amplitude (log) | 0.43 - [0.29; 0.56] | 0.94 - [0.90;0.96] | 0.16 |
| Total tremor acceleration (log) | 0.54 - [0.4; 0.64] | 0.93 - [0.87;0.96] | 0.16 |

**Supplementary Table 3.** Postural tremor methods.

### Postural tremor

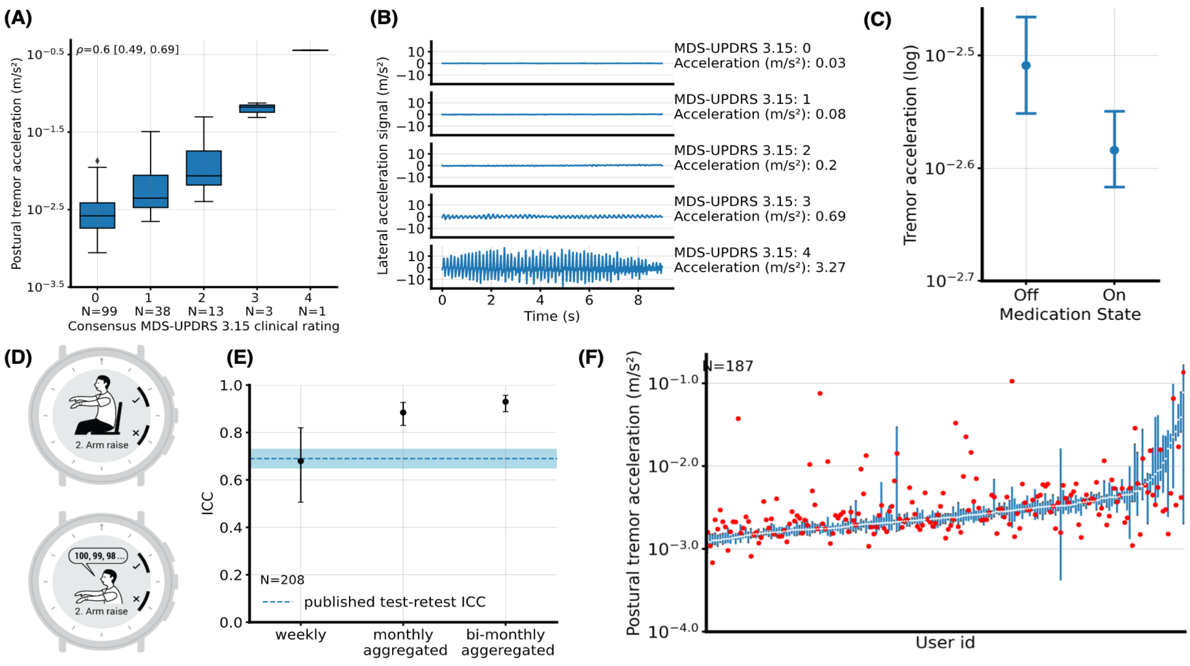


**Supplementary Fig 1. Postural tremor acceleration (A)** Lateral tremor acceleration (log scale) measured during the in-clinic examination, separated by postural tremor (MDS-UPDRS 3.15) consensus scores. **(B)** Raw acceleration signals, along the y axis, collected from example participants, are associated with each clinical score. Measurement values, as computed by the PD-VME are also indicated. **(C)** Mean and 95% Confidence interval of at-home lateral tremor acceleration, aggregated over a 2-month baseline period, in two different medication states: On and Off-levodopa. **(D)** User interface providing participant-facing instructions when performing the unsupervised virtual tremor assessment. **(E)** Intra-class correlation between at-home measurements. Whiskers represent 95% confidence intervals. The first column measures week-over-week test-retest reliability of a single measurement. The second column measures the month-over-month test-retest reliability of the monthly-averaged measurements. The third column is similar but with bi-monthly averaging. The dotted blue line and light blue shading represent the published postural tremor test-retest ICC[1] and the associated 95% confidence interval. **(F)** Distribution of tremor measurements in the Off-medication state, by participant. Participants are ordered according to their median at-home PD-VME measurement. Each vertical blue bar represents the 25th and 75th percentile of at-home measurements for a given participant. Red dots represent the in-clinic sensor measurement obtained during the MDS-UPDRS assessment.

The method to compute postural tremor acceleration was chosen to match the one selected for rest tremor. The rest of the analysis follows the same logic.

The Spearman rank correlation between the log of the median wrist acceleration during the postural tremor task and expert consensus rating of MDS-UPDRS task 3.15 was $\rho = 0.60$ $[0.49, 0.69]$, $N = 154$ (Fig. 4.A). Change (Cohen's D of 0.19) was observed in the tremor acceleration when comparing before and after levodopa intake. (Fig. 4.C). Week-on-week intra-class correlation (ICC) of 0.68 [0.51-0.82] is shown in figure 4.E (N=208). When the measurements are averaged over one month, the month-on-month test-retest ICC increased to 0.90 [0.83-0.95] (N=139).

### Bradykinesia methods

The PD-VME task to measure bradykinesia resembles the MDS-UPDRS 3.6 pronation-supination task: participants are asked to twist their arms, hands extended, as widely as possible, for a duration of 20 seconds.

Two methods were considered for bradykinesia estimation: arm twist amplitude (in degrees), and arm rotational speed (in degrees per second).[2] For both methods, correlation against the in-clinic MDS-UPDRS pronation-supination (3.6) single-rater score was similar, and arm twist amplitude was selected. Arm twist amplitude was computed along the x-axis of the gyroscope sensor on Study Watch by integrating the signal using polynomial detrending and taking the median value.

To more precisely detect the start and end of the task, a segmentation method was applied by selecting the first and last times where the gyroscope signal exceeds a given threshold. Thresholds were selected out of a total of 6 combinations, based on the correlation to single-rater MDS-UPDRS scores.

| Method | Test-retest ICC (week-on-week) | On/Off effect size | Spearman R vs clinical ratings |
| --- | --- | --- | --- |
| Arm twist amplitude (degrees) | 0.71 | -0.07 | -0.62 |

| | | | |
|---|---|---|---|
| Arm twist rate (Hz) | 0.77 | -0.22 | -0.06 |
| Arm twist speed (degrees / s) | 0.75 | -0.27 | -0.26 |
| Arm twist combination | 0.77 | -0.26 | -0.41 |

**Supplementary Table 4.** Bradykinesia methods.

#### Gait methods

The PD-VME task for gait task resembles a combination of the MDS-UPDRS 3.9 sit-to-stand and 3.10 gait tasks. Participants are instructed to get up from a chair, with arms crossed over their chest, and walk back and forth for a duration of 60 seconds.

Several methods[3-5]were considered to quantify gait impairment symptoms. The arm swing amplitude[5] was explored owing to high correlation with single-rater MDS-UPDRS 3.10 ratings, and its published ability to capture disease progression, and act as a prodromal marker of PD.[6–8] Arm swing acceleration captures the maximum range of vertical acceleration of a PD patient while they are walking. It is defined as the norm of the x-axis and y-axis accelerations. Additionally, arm swing forward acceleration was calculated and compared. This feature captures the load changes of the wrist acceleration from the repetitive impacts from steps during walking. It is defined as the rate of change of forward acceleration (x-axis) during walking.[3] Spectral features extracted from the power spectral density (PSD) within the walking frequency (<2.5 Hz) were also explored. These features are the peak frequency in the PSD and the corresponding peak magnitude. Peak frequency is defined as the cadence (i.e., the walking speed) during the MDS-UPDRS walking task. The peak arm swing energy was also calculated, which is defined as the peak magnitude within the walking frequency (<2.5 Hz) in the power spectral density (PSD).

| method | R (spearman) vs clinical scores | Test-retest ICC (month-on-month) | on_off (Cohen's D) |
|---|---|---|---|
| Arm swing acceleration | -0.46 - [-0.58; -0.31] | 0.75 - [0.659;0.831] | -0.45 |
| Peak arm swing energy | -0.49 - [-0.59; -0.37] | 0.82 - [0.686;0.91] | -0.31 |

| | | | |
|---|---|---|---|
| Arm swing forward acceleration | -0.37 - [-0.5; -0.24] | 0.89 - [0.819;0.936] | -0.54 |
| Cadence | -0.1 - [-0.26; 0.07] | 0.76 - [0.669;0.842] | -0.17 |

**Supplementary Table 5.** Gait methods.

### Relationship of unsupervised sensor measurements when collected in the clinic and at home

Besides comparing supervised in-clinic measurement with the unsupervised in-clinic measurement we extended the analysis to look for systematic differences in the unsupervised measurements collected in the home and in the clinic.

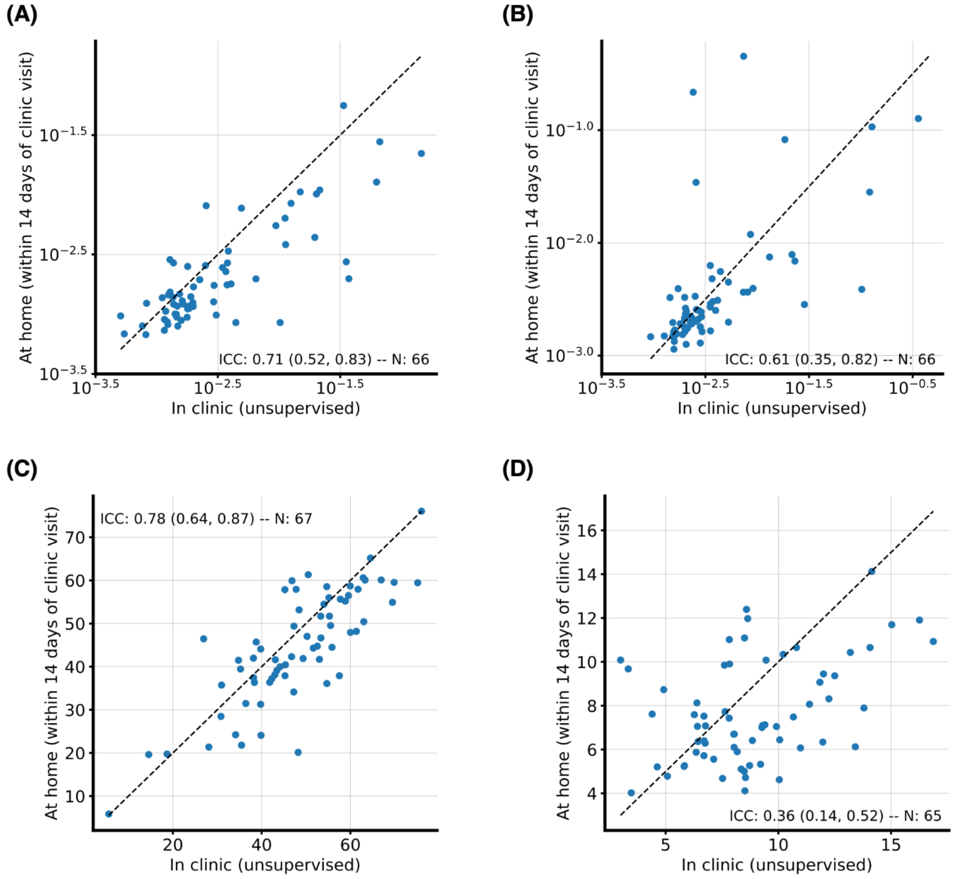

**Supplementary Fig 2. Comparison of the unsupervised measurements taken in-clinic and at-home** within 14 days, in the OFF-medication state. (A) Rest tremor acceleration. (B) Postural tremor acceleration. (C) Arm twist amplitude. (D) Arm swing acceleration.

In supplementary Fig 2. (A-D), we compare the unsupervised measurements taken in-clinic and at-home within 14 days, in the OFF-medication state. We find ICC=0.71 [0.52-0.83] and ICC=0.61 [0.35-0.82] for the rest and postural tremor accelerations respectively, and ICC=0.78 [0.64-0.87], and ICC=0.36 [0.14-0.52] for the arm twist amplitude and arm swing acceleration during gait respectively.
